# Synchronizability predicts effective responsive neurostimulation for epilepsy prior to treatment

**DOI:** 10.1101/2021.02.05.21250075

**Authors:** Brittany H. Scheid, John M. Bernabei, Ankit N. Khambhati, Jay Jeschke, Danielle S. Bassett, Danielle Becker, Kathryn A. Davis, Tim Lucas, Werner Doyle, Edward F. Chang, Daniel Friedman, Vikram R. Rao, Brian Litt

**Author notes:** Correspondence to: Brittany H. Scheid, Department of Bioengineering, University of Pennsylvania, Philadelphia PA 19104 USA.

## Abstract

Despite the success of responsive neurostimulation (RNS) for epilepsy, clinical outcomes vary significantly and are hard to predict. The ability to forecast clinical response to RNS therapy before device implantation would improve patient selection for RNS surgery and could prevent a costly and ineffective intervention. Determining and validating biomarkers predictive of RNS response is difficult, however, due to the heterogeneity of the RNS patient population and clinical procedures; large, multi-center datasets are needed to quantify patient variability and to account for stereotypy in the treatment paradigm of any one center. Here we use a distributed, cloud-based pipeline to analyze a federated dataset of intracranial EEG recordings, collected prior to RNS surgery, from a retrospective cohort of 30 patients across three major epilepsy centers. Based on recent work modelling the controllability of distributed brain networks, we hypothesize that broader brain network connectivity, beyond the seizure onset zone, can predict RNS response. We demonstrate how intracranial EEG recordings can be leveraged through network analysis to uncover biomarkers that predict response to RNS therapy. Our findings suggest that peri-ictal changes in synchronizability, a global network metric shown to accurately predict outcome from resective epilepsy surgery, can distinguish between good and poor RNS responders under the current RNS therapy guidelines (area under the receiver operating characteristic curve of 0.75). Furthermore, this study also provides a proof-of-concept roadmap for multicenter collaboration where practical considerations impede sharing datasets fully across centers.

## Introduction

Approximately 1 in 26 people will develop epilepsy at some point in their lifetime, and nearly one third of epilepsy patients are medication-resistant and experience recurrent seizures. Although surgically removing the seizure onset zone (SOZ) holds the highest promise for medication-resistant patients to become seizure-free, resective surgery is often not a viable option for patients with multiple seizure onset zones, seizures that originate from eloquent cortex, or a spatially extensive seizure onset.^1^ Responsive neurostimulation (RNS) therapy offers a promising alternative to traditional resective surgery for these patients.^2^ The current FDA-approved RNS device for epilepsy consists of an implantable pulse generator affixed to the skull connected to two subdural electrodes that facilitate continuous intracranial EEG (iEEG) sensing and direct electrical stimulation when abnormal activity is detected.^3^ A recent review of RNS outcomes in a nine-year prospective follow-up study showed that over 50% of patients are RNS responders (≥ 50% reduction in seizures) after one year, and over 70% of patients are responders at three years^4^.

Although RNS is effective for many patients, approximately 10% of patients demonstrated no change or an increase in seizure frequency after nine years of RNS therapy, and less than 20% of patients experienced seizure freedom continuously for a period greater than one year.^4^ The RNS implantation procedure is highly invasive and costly, and the risk of serious adverse events including tissue infection, osteomyelitis, and hemorrhage, while small, is especially burdensome for patients whose condition is not improved by the device.^5–7^ Additionally, candidates for RNS typically have a multi-year history of failed medications and may have failed surgical interventions^2,8^; the psychological toll of prolonged treatment failure can exacerbate comorbid anxiety and depression that are prevalent in the refractory epilepsy population.^9,10^ An expanding body of work is uncovering clues to the mechanism of RNS action^2,11–15^, but there is an ongoing need for validated biomarkers to improve RNS therapy. In particular, a biomarker that indicates whether RNS therapy will lead to a successful outcome, given that the current therapeutic guidelines for RNS therapy are followed^16^, and that is measured *before* device implantation could be a critical tool for physicians to consider when weighing therapeutic options for their patients.

To date, many of the biomarkers associated with RNS patient outcomes, such as interictal spike frequency and spontaneous seizure interruption, have been extracted from long-term RNS device recordings after device implantation.^12,17^ These studies suggest that progressive restructuring of the epileptic network due to chronic RNS action, rather than acute, focal seizure suppression by electrical stimulation, has the greatest impact on reduced seizure frequency in responders. Data collected in the epilepsy monitoring unit (EMU) during presurgical monitoring remains an unexplored resource for guiding RNS placement beyond localizing the seizure onset zone, and holds promise for discovering RNS biomarkers that can predict patient outcome. iEEG recordings in particular have been used to predict outcomes from resective surgery^18,19^, indicate effective locations and time points for seizure control^20^, predict dynamics of seizure spread^21^, and functionally map brain networks through corticocortical evoked potentials.^22,23^ It is likely that multiple factors, such as medication schedule, RNS implant location, and stimulation parameters contribute to a patient’s therapeutic response. Still, evidence that RNS therapy has a gradual effect leads to the hypothesis that the organization of brain network connectivity may help predict response to targeted RNS therapy.

In this study, we present a candidate biomarker— network synchronizability— calculated from iEEG recorded during presurgical evaluation, for predicting whether a patient will respond to RNS therapy. Network synchronizability is a metric borrowed from the field of graph theory and is a theoretical measure of the diffusion of information throughout a network under certain assumptions of system dynamics.^24,25^ Preliminary studies show that the change in network synchronizability at seizure onset is a promising candidate biomarker for identifying patients most likely to benefit from resective surgery.^18,26^ In the context of RNS therapy, multiple theories surrounding the effect of RNS on synchronization within the epileptic network suggest that the functional connectivity of brain networks during seizures may also be useful in distinguishing RNS responders from non-responders. Kokkinos *et al*.^12^ found that the dominant power spectral frequency at the beginning of an electrographic seizure evolves over chronic RNS therapy, suggesting that effective therapy promotes desynchronization among brain regions involved in seizures. Therefore, patients whose epileptic network is more amenable to perturbation by RNS therapy may exhibit better clinical outcomes. In this study, we test the hypothesis that peri-ictal global brain network iEEG dynamics recorded before RNS implant can indicate whether a patient will respond to RNS therapy. We also we establish a set of shared inclusion criteria, computational pipelines, and data repositories to analyze intracranial electrophysiology and clinical metadata in a federated manner across multiple epilepsy centers. We anticipate our federated framework to facilitate the evaluation of candidate pre-implant biomarkers in the future.

## Methods

### Patient Selection and Data Collection

We aggregated a retrospective cohort of 30 patients, who had been implanted with an RNS device between June 2014 and September 2018, across three major epilepsy centers— ten patients per center. We included patients who were over 18 years of age, underwent at least two years of RNS treatment, experienced at least one seizure per month before RNS treatment, and underwent iEEG monitoring during pre-surgical evaluation in the epilepsy monitoring unit (EMU) of their RNS treatment center as part of their standard clinical care before RNS implant. Patients who underwent resective surgery concurrent with RNS therapy were excluded from this study. Ten out of 19 consenting patients from the Hospital of the University of Pennsylvania (HUP) met the inclusion and exclusion criteria at the time of data assembly, and were selected for this study. Ten patients meeting the inclusion and exclusion criteria at New York University (NYU) Langone Comprehensive Epilepsy Center and University of California, San Francisco (UCSF) Comprehensive Epilepsy Center, respectively, were additionally selected as a convenience sample to build a 30-patient dataset with equal representation from each center (**Table S1)**. iEEG data was recorded from cortical grids and strips, depth electrodes, stereo EEG (SEEG) electrodes, or a mixture of electrode types across centers, sampled at either 500 Hz, 512 Hz or 1024 Hz (with one exception-recordings of patient NP47 were down sampled from an 8192 Hz sampling rate to a 1024 Hz sampling rate before further processing). At each center, iEEG signals were recorded referentially, with the reference electrode placed distant to the site of seizure onset. Patients were determined to be good candidates for the NeuroPace RNS^®^ system based on consensus of each center’s multidisciplinary epilepsy care team during epilepsy surgical conference. Data from these evaluations were collected solely for clinical use and incorporated into this study retrospectively. Data collection for research purposes at HUP was approved by the HUP Institutional Review Board under the Collaborative and IEEG protocols; all HUP subjects provided consent to have their full-length intracranial EEG recordings and anonymized imaging and metadata publicly released on the ieeg.org portal, an open-source online repository for electrophysiologic studies. The NYU Langone Institutional Review Board granted approval for data collection and allowed that informed consent could be waived for studies involving sharing of deidentified EEG and imaging data. Finally, data collection at UCSF was approved by the UCSF Committee on Human Research, which ruled that informed consent could be waived for studies involving sharing of deidentified EEG and imaging data.

We quantified a patient’s response to RNS treatment as the percent change in seizure frequency at their most recent clinical visit compared to their pre-implant baseline frequency, as reported in the patient’s seizure diary and clinical notes. We classified patients that achieved ≥ 50% seizure reduction as ‘responders’ (N=19), a standard threshold for treatment evaluation, while those who had a lesser reduction or increase in seizures were classified as ‘non-responders’ (N=11).^6^

### Intracranial EEG Processing & Functional Network Generation

Seizures were identified in the iEEG recordings at each center during clinical evaluation, and the seizure onset and seizure termination timepoints were annotated by board-certified epileptologists (BL, VRR, DF).^27^ In total, 159 seizures across 30 patients were identified for analysis (median: 3 seizures/subject, IQR: 2 – 6). Artifactual channels were identified by visual inspection and removed. Next, data clips containing an equal period of preictal recording preceding each seizure followed by the seizure itself (median seizure length: 1.4 min, IQR: 45 sec – 2.7 min), were formatted to enter the pre-processing and network generation pipeline described in previous publications.^18,26^ Briefly, the raw iEEG recordings were de-noised using a common average referencing (CAR) technique, wherein the mean across recording channels at a given time point is subtracted from all channels at the same time point.^26,28^ Importantly, CAR is robust to the placement of the reference electrode used for signal recording, thus mitigating one potential difference in recording technique between centers.

Metrics from network theory can be applied to electrographic neural data by constructing functional connectivity networks in consecutive time windows, using iEEG electrodes as nodes and assigning the strength of coherence between pairs of electrode recordings as weighted edges. Accordingly, each event clip was split into a total of 2*T* non-overlapping 1-second time windows (*T* preictal, *T* ictal windows), and functional networks were generated for each time window such that the *N* recording electrodes represented *N* network nodes, and an estimate of coherence between each pair of 1-second channel recordings represented the edge weight between respective electrode pairs. We used multitaper coherence, a measure of similarity in spectral power between two signals at a given frequency, to calculate network edge weights as the average coherence value across frequencies in a given frequency band. Studies of neural communication in a variety of behavioral contexts find that neuronal coherence may increase within distinct frequency bands to achieve specific neurophysiological aims, with feedforward communication signals mediated by higher frequencies and feedback signals mediated by lower frequencies.^29^ Thus, coherence networks built using specific frequency bands can tease apart band-specific dynamics of neural communication. Therefore, a multitaper coherence network was generated over the β band (15-25 Hz) and high-γ band (95-105 Hz) respectively, as well as a cross-correlation network calculated over a broad band range (5-115 Hz) for each time window.^18^ We were left with three functional connectivity networks with *N* nodes for each of the *T* time windows preceding and *T* time windows following seizure onset for each seizure (**Fig. 1A**).

**Figure 1.**
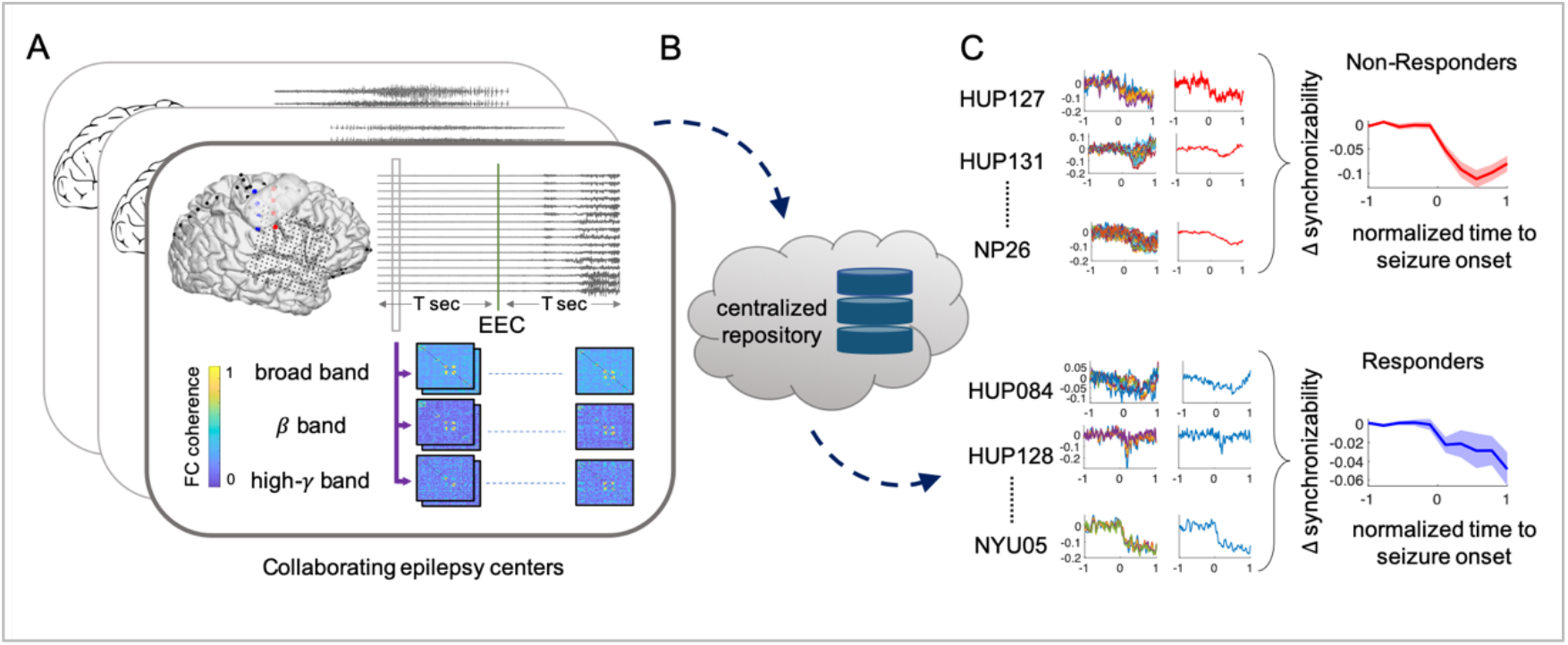
Construction of functional networks and synchronizability curves. (A) Functional connectivity networks, represented by square adjacency matrices, are generated from consecutive 1-second windows in the pre-ictal and ictal periods of each seizure event. The same processing pipeline, represented by purple arrows, is used within the data environment of each institution to estimate multi-taper coherence networks in the β band and high-γ bands, as well as broad band cross-correlation networks. (B) All processed networks are shared in a centralized cloud repository. (C) Synchronizability curves are calculated for each seizure, re-referenced to a pre-ictal baseline, normalized to a unit length, and averaged over 10 time bins within a patient. Patient curves are grouped by responder status and averaged to yield a representative responder curve. EEC-earliest electrographic onset; FC-functional connectivity.

### Network Synchronizability

The metric of network synchronizability is a global network metric that estimates the ease of information propagation throughout a functional connectivity network^30^. We calculate the synchronizability value for consecutive functional connectivity networks to create a synchronizability curve as a function of time, s(t). Synchronizability for a given time window is calculated by first representing a network as an N x N adjacency matrix, ***A***, where the *ij*)* element holds the value of the edge weight between nodes *i* and *j*. The Laplacian matrix is then calculated as ***L*** = ***D*** − ***A***, where ***D*** is a diagonal matrix of node strength.^24^ Finally, synchronizability is given as the ratio of the second smallest to the largest eigenvalue of the Laplacian matrix.

Once we obtain a synchronizability curve for each seizure event, we re-reference the curves by the mean synchronizability value over the first half of the pre-ictal period. Thus, the amplitude of the shifted curves represents the change in synchronizability from a pre-seizure baseline. Each synchronizability curve is normalized such that the ictal period has unit length and the curve is segmented into 10 equally spaced time bins (5 pre-ictal, 5 ictal), with a given bin value calculated as the average of synchronizability values in the associated time interval. For each patient with multiple seizures, all 10-point shifted synchronizability curves are averaged across each time bin, resulting in a single representative curve per patient, used for group level analysis (**Fig. 1C**).

### Aggregating Data in a Federated Framework

We created a framework that allowed for distributed processing of clinical neuromonitoring data across epilepsy centers in a standardized manner. Ten patients were selected using the same exclusion and inclusion criteria at each center, respectively, and gave their written informed consent to share their de-identified neuromonitoring data. Mutual data-use agreements were prepared between each institution to allow the investigators to share limited datasets and post-processed results on the International Epilepsy Electrophysiology Portal (ieeg.org)^31^, a centralized HIPPA-compliant cloud repository. The same pipeline for electrophysiological data pre-processing and network generation was distributed across centers^26^, ensuring that the data processing steps were identical across sites.

### Statistical Analysis

We used the curve comparison test from functional data analysis (FDA)^32^ to compare the areas under the averaged responder and non-responder synchronizability curves throughout pre-seizure and seizure periods. We also performed a sensitivity analysis to quantify how well the change in synchronizability predicted the patient response. We generated a receiver operating characteristic (ROC) curve to measure how well the change in synchronizability for a given patient, measured as the median synchronizability value in the first half of the pre-ictal period subtracted from the median synchronizability value of the ictal period, could classify a patient as a responder or a non-responder for a sweep of classification thresholds. The area under the ROC curve (AUC) was measured, with a value of one representing perfect classification, and a value of 0.5 representing chance assignment of a patient to one of the two groups.

### Data availability

We share all functional connectivity networks and synchronizability curves derived from iEEG recordings obtained during a patient’s stay in the epilepsy monitoring unit on the free and HIPPA-secure web portal ieeg.org.^31^ The code used for generating functional networks and calculating synchronizability is freely available at https://github.com/akhambhati/Echobase.

## Results

We began our analysis by examining whether quantifiable clinical factors alone were associated with patients’ RNS response. We found no significant difference between responder and non-responder groups based on years of RNS therapy (responder IQR= 2.9 − 4.0, non-responder IQR= 3.0 − 3.6, Mann-Whitney *U*= 302.5, *p* > 0.7), years with epilepsy (responder IQR= 13 − 35.8, non-responder IQR= 14 − 28.5, Mann-Whitney *U*= 295, *p* = 1), or the location of RNS lead implants (mesial temporal implant *versus* neocortical, Mann-Whitney *U*= 229, *p* > 0.6, unilateral *versus* bilateral *U*= 239, *p* > 0.9). Additionally, there was no significant difference in number of implanted iEEG electrodes between groups (responder IQR= 97 − 149, non-responder IQR= 86 − 137, *U*= 300, *p* > 0.8). Given that clinical features did not distinguish responders from non-responders, we next turned to our hypothesis that synchronizability, a measure based on the connectivity of functional brain networks, might have value for predicting treatment response.

The synchronizability value measured from functional brain networks, built using a measure of neural coherence between macroscopic brain regions, estimates the capacity for neural information to diffuse throughout a network.^26,30,33^ Thus a change synchronizability after seizure onset is a measure of how much the neural channels for communication are impeded or facilitated during the seizure, compared to a baseline synchronizability value. We quantified how the change in synchronizability at seizure onset differed between RNS responders (patients showing at least a 50% reduction in seizure frequency compared with baseline) or non-responders (all other patients) at their most recent visit after at least 2 years of RNS titration. We computed synchronizability changes in two frequency bands, *β* (15-25 Hz) and high-*γ* (95-105 Hz), and broad band (5-115 Hz), and found that RNS responders demonstrated a smaller decrease in network synchronizability after seizure onset compared with patients who were non-responders. The difference was significant in the broad band (*p* < 0.001) and the *β*-band (*p* < 0.002) using an uncorrected FDA test for statistical significance. Even after using a Bonferroni correction for multiple comparisons across the three distinct network types, the broad band and *β*-band maintained significance (*p* < 0.0167) The decrease in synchronizability was similar for both groups in the high-γ band (*p* > 0.05), suggesting that this frequency band does not contain network information pertinent to predicting patient response (**Fig. 2**). In a sensitivity analysis, synchronizability change for *β*-band and broad band networks was able to predict responder outcome with an AUC value of 0.75 for broad band and an AUC value of 0.72 for *β*-band data (**Fig. 3**).

**Figure 2.**
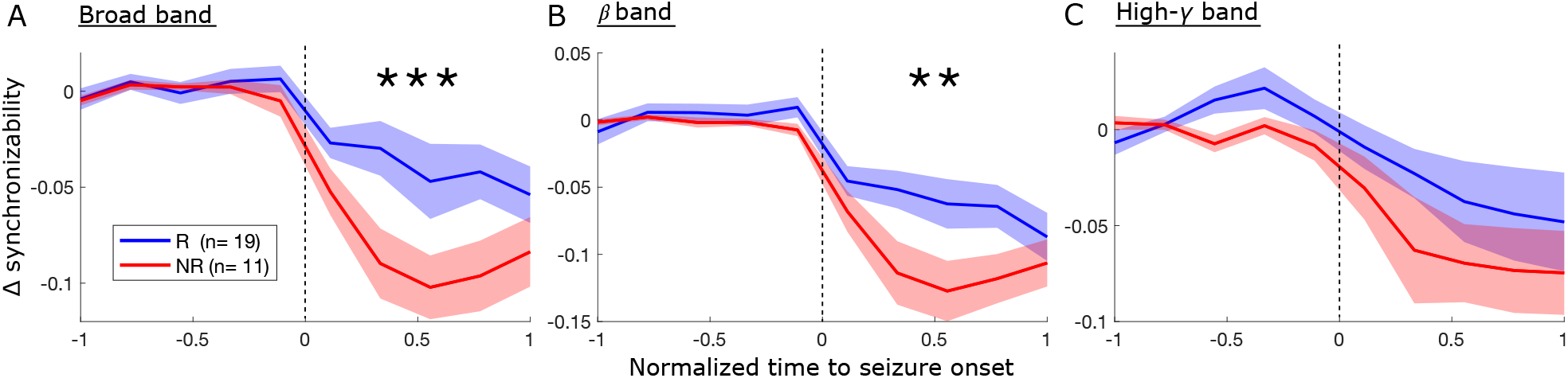
Time-varying synchronizability separates RNS responders from non-responders. We generated functional coherence networks from sequential time windows of iEEG seizure recordings from 30 RNS patients. We then calculated the synchronizability of each network to obtain a time varying synchronizability curve per seizure. All synchronizability curves were normalized to unit length pre-ictal and ictal periods. The curves were averaged across seizures in a given subject and discretized into 10 uniform time bins. Solid lines in the plots show the averaged synchronizability curves of subjects sharing the same responder status (R-responder, NR-non-responder). The envelope indicates the standard error of the mean. Synchronizability decreased significantly in non-responders after seizure onset compared to responders for curves calculated using broad band (5-115 Hz) coherence networks (N=30, p<0.001) (A), and β-band (15-25 Hz) coherence networks (N= 30, p<0.002) (B), but not for high-γ (95-105 Hz) coherence networks (N=30, p > 0.05) (C). Significance was calculated using functional data analysis, with p<0.05 deemed significant.

**Figure 3.**
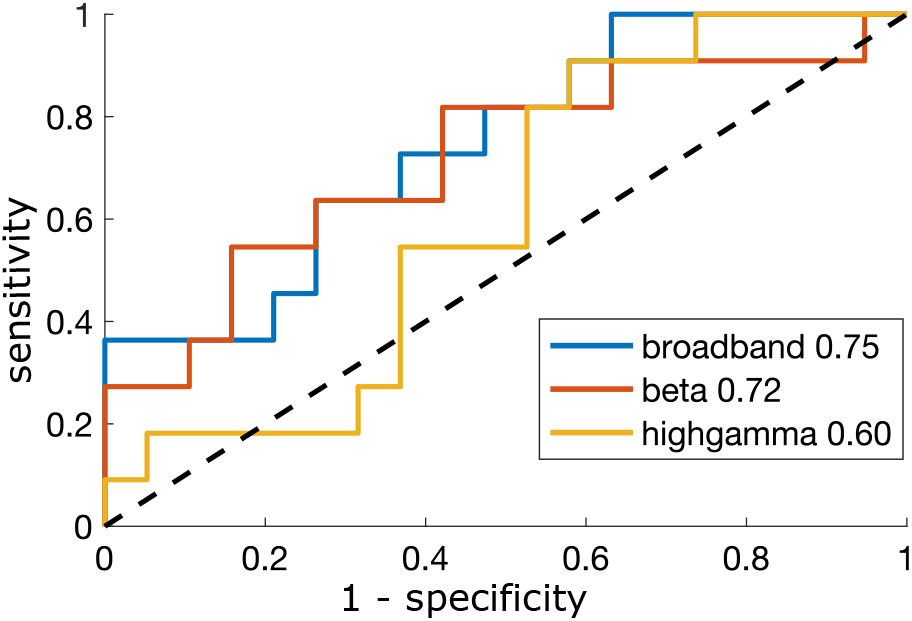
ROC curves for synchronizability as a predictor of RNS outcome. The change in synchronizability for a given patient was calculated as the median synchronizability value in the first half of the pre-ictal period subtracted from the median synchronizability value of the ictal period. ROC curves show the true positive rate (sensitivity) versus the false positive rate (1-specificity) at each classification threshold. Data were generated for the two frequency bands and broad band. The area under the ROC curve (AUC) was greatest for broad band networks with an AUC of 0.75.

We next performed two sub-analyses to determine whether the main effect of a greater synchronizability decrease at seizure onset in non-responders held after patients were grouped by iEEG electrode type and by location of implanted RNS leads. The difference between the median synchronizability value in the first half of the interictal period and the median synchronizability value in the ictal period was quantified for each patient. We found that change in synchronizability was significantly different between responders and non-responders in the broad band for patients with bilateral implantation of RNS leads (*nR* = 8 *nNR* = 3, *U* = 58, *p* < 0.05). In general, the trend of a greater synchronizability decrease in non-responders was maintained in each category for broad band and *β* band synchronizability measures, but varied across high-*γ* band synchronizability (**Fig. 4**). An additional analysis on patients within each center also demonstrated trends consistent with the main findings in the broad band and *β* band, though the individual center results did not reach significance (**Supplementary Fig. 1**). The results illustrate that neither laterality of implant, type of intracranial electrodes, nor treatment center bias the main findings of responder differences found for broad band and *β* band synchronizability change.

**Figure 4.**
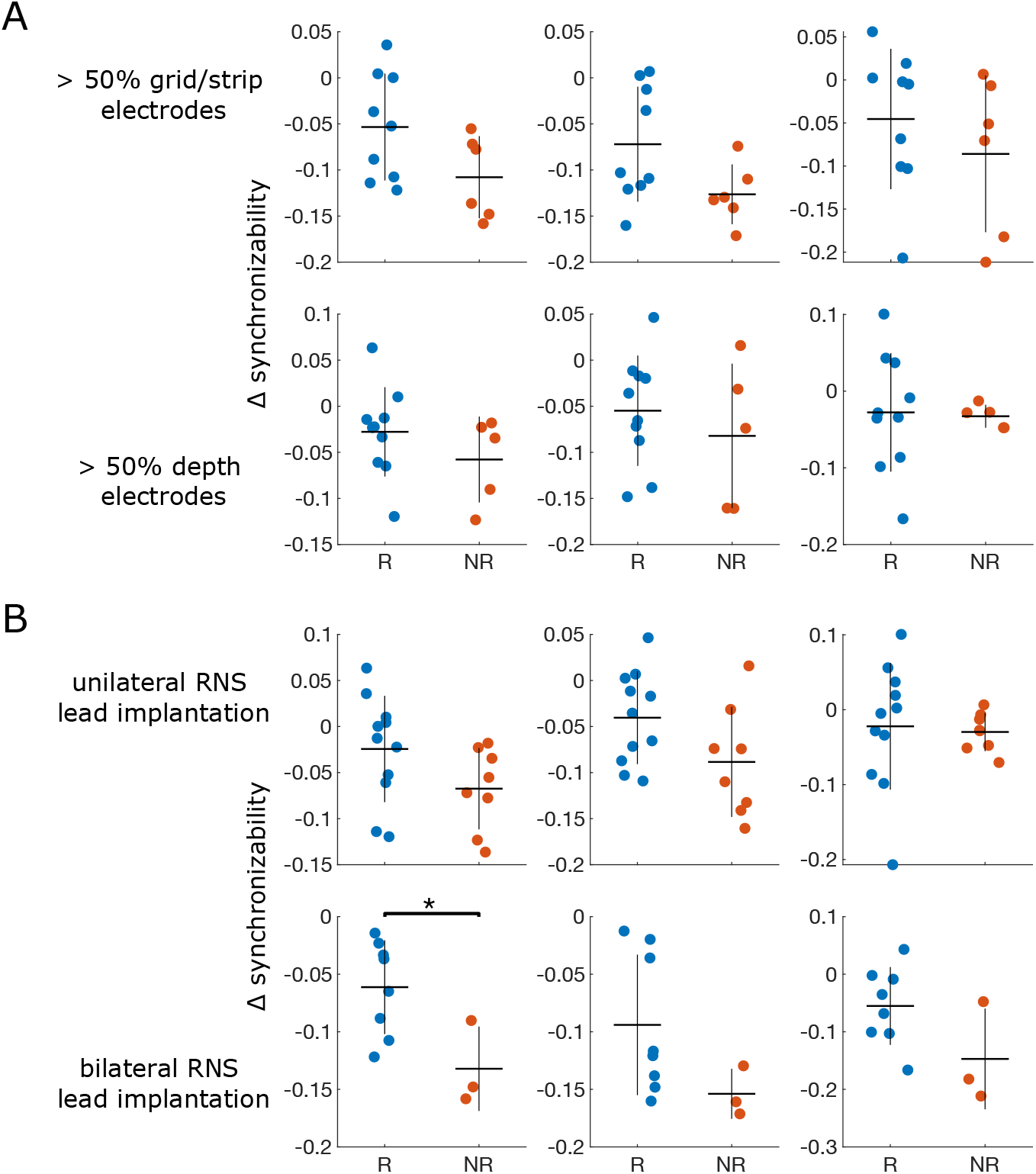
Synchronizability changes among patient subgroups. Separation of responders and non-responders in each synchronizability band for patients with < 50% grid/strip electrodes vs. patients with majority depth electrodes (A), and in patients with unilateral vs. bilateral onset foci (B). Horizontal lines represent the mean synchronizability change within a response group (R-responders, NR-non-responders), the vertical lines represent the standard deviation within a group.

## Discussion

To our knowledge, our results are the first to demonstrate the prognostic value of iEEG data collected before RNS implantation to determine whether patients are likely to respond to RNS therapy. In our analysis, we find that the biomarker of synchronizability change at seizure onset has the ability to distinguish current RNS responders from non-responders, with trends maintained even after segmenting patients by onset location, iEEG electrode type, and treatment center. Additionally, our study serves as a proof-of-concept for multi-center collaborative RNS biomarker discovery on federated datasets.

Much evidence suggests that epilepsy is a disorder of brain networks^34^, and incorporating measures of complex macroscale neural dynamics is promising for guiding surgical resection in cases of medically refractory epilepsy.^35,36^ In prior work, a decrease in the global network measure of synchronizability at seizure onset predicted a good surgical resection outcome, reflecting the network’s ability to isolate seizure propagation at seizure onset.^18^ Our finding that a large decrease in synchronizability at seizure onset is associated with poor RNS outcomes suggests that neurostimulation may operate via a fundamentally different mechanism than resective surgery, and that unique neurophysiological features can portend favorable prognosis with neurostimulation compared to resection. Analysis of longitudinal RNS recordings has led to the theory that stimulation in RNS responders may actuate long-term plasticity changes to fragment tightly-coupled groups of neurons that would otherwise synchronize through seizure onset.^12,14^ A purely speculative possibility is that by maintaining a similar level of synchronizability between pre-ictal and ictal periods as shown in our broad band results, RNS responders may be better able to distribute energetic inputs to the broader epileptic network during seizures, when the device is commonly triggered for stimulation. In patients with decreased post-onset synchronizability, RNS may be less able to influence long-range plasticity mechanisms in the broader epileptic network.

The mechanism of RNS remains poorly understood, and undoubtedly a number of factors may influence responder outcome, including location of RNS implant with respect to a patient’s seizure onset zone^37^, programming detection and stimulation parameters^13^, and interactions between RNS therapy and pharmaceutical treatments.^38^ Much of the RNS parameter space has not yet been explored, and it is unclear to what extent each factor may contribute to patient outcome. Thus our results suggest that the inherent connectivity within a patient’s epileptic network may be a significant predictive factor of their RNS response, given the current RNS treatment guidelines. The fact that our results are not sensitive to unilateral *versus* bilateral RNS lead placement is consistent with clinical data that finds similar reductions in seizure frequency regardless of RNS lead location.^4^ Taken together, our study extends a growing body of literature suggesting that RNS action may be a global network phenomenon, and that chronic plasticity changes may be a relevant factor in RNS efficacy, beyond the mechanism of acute seizure termination for which RNS was originally designed.^12^

Using intracranial EEG during presurgical evaluation to determine if a patient should be implanted with an RNS device is not a novel concept. Patients are typically referred for RNS therapy after iEEG implant when seizures are found to emanate from an eloquent region that cannot be resected, when there are multiple, spatially distinct generators for seizures (e.g. independent bitemporal seizure onsets), or when seizures are poorly localized and focal resection or ablation is not an option.^2^ It is typically presumed that in the first two situations RNS is more likely to be effective, though we know that rapidly synchronized networks may exist in well-localized focal or multifocal epilepsies, likely related to their underlying cause. It is our long-term-vision that our biomarker may identify these poor responders early, and that it may also identify potential good responders whose seizures spread rapidly or are poorly localized. In our study there were no features in the iEEG seizure onset pattern that distinguished likely responders from non-responders, even on review by expert epileptologists. We do not yet know whether the utility of peri-ictal network synchronizability as a biomarker of treatment response is unique to RNS or is a general marker of the susceptibility of the epileptic network to neuromodulation including deep brain stimulation and vagus nerve stimulation. Rigorous quantitative analysis of many more patients will be required to more precisely understand the physiologic underpinnings of our predictions.

### Building a multi-center collaborative pipeline

Procedures for pre-implant evaluation and RNS treatment, including patient selection, lead placement, and device programing, are not standardized across epilepsy centers and differ from protocols used in RNS clinical trials.^16^ Thus, results from clinical trials will need to be augmented by quantitative biomarkers, not just to select patients for RNS therapy, but also to select regions for stimulation and to titrate stimulation parameters. Although candidate biomarkers to predict RNS response exist, discovering robust biomarkers and validating them is challenging for a number of reasons. First, the numerous causes of epilepsy contribute to a heterogeneous patient population compared with other disorders like Parkinson’s disease, where neural pathways and stimulation targets are well-characterized, and stimulation parameters can be rapidly tuned based on symptom abatement^39,40^. Due to heterogeneity of epilepsy subpopulations, it is likely that RNS patients will require precise therapeutic approaches that depend on their etiology and underlying network dysfunction^41^. Second, the hypothesis and parameter space to consider for RNS therapy is large, encompassing questions of electrode positioning, stimulation parameters, detection thresholds, and timing. Finally, although a centralized repository of RNS device data exists with the device manufacturer, the manufacturer does not have access to clinical details about changes in patients’ medication, behavior, or outcome.

Many of the barriers to biomarker discovery in general can be mitigated by assembling datasets that are large enough to yield required statistical power for validation among specific clinical subgroups, however in the case of RNS studies there are currently no individual clinical sites that have both the engineering infrastructure and the hundreds of implanted RNS patients necessary for the investigations outlined in this study. In an age of digital health records and secure cloud-based platforms for data processing, multicenter collaborations are an effective way to quickly build powerful datasets, so long differences in data collection and processing techniques between centers are accounted for. Positive findings and hypotheses can be tested and cross-validated using data from multiple centers, ensuring that results are unbiased and robust to variability in clinical protocols among sites. Still, there are real obstacles to sharing identifiable data, such as ensuring patient privacy and adherence to IRB protocols, and sharing may also be limited when working with industry partners who may be bound by regulatory guidelines and other practical constraints on data distribution.

For these reasons we elected to unify data formats, annotation protocols, electrode coregistration and analysis code so that our experiments could be federated, or performed separately and in parallel across our sites, and then the results aggregated centrally. Despite some initial up-front effort to make this protocol run smoothly, we found this approach to be very feasible and efficient, once the work commenced. In this way, our study lays the groundwork for streamlined collaboration on RNS biomarker discovery. We only discovered the significant differences in synchronizability change based on RNS outcome after combining data across multiple centers; a result that is a powerful demonstration of how biomarker evaluation using any one institution’s dataset is statistically underpowered, and shows that only by aggregating data for analysis can the potential biomarker be seriously assessed. We believe that this same paradigm could be extremely useful in other difficult “medical informatics” problems that require analyzing large amounts of data across institutional and industry boundaries. By utilizing centralized tools that can be ported into center-specific data environments, investigators can uphold privacy and firewall restrictions on original data while sharing their processed derivatives.

### Methodological Considerations

One clinical challenge presented in the study of RNS patients involves assessing clinical outcome. While in resective and ablative surgery there are standardized ILAE and Engel outcome scales, the majority of patients undergoing RNS do not experience complete seizure freedom, and success is gauged by seizure reduction. The RNS device quantifies electrographic seizures through counts of ‘long episodes’, sustained detections of epileptiform activity that exceed a pre-specified duration. However, long episodes are not a reliable proxy for electrographic seizures in all individuals; long episodes can reflect prolonged stretches of abundant interictal discharges that meet detection criteria but are not sufficiently organized or temporally evolving to constitute seizures.^42^ While steps have been taken to estimate the positive predictive value for a long episode representing an electrographic seizure^43^, the sensitivity of the device can be adjusted, so there is not a reliable way to determine whether all seizures are represented. For this pilot study we chose to use each patient’s self-reported seizure diary, which is the currently accepted gold-standard for calibrating RNS therapy, as our measure of therapy response. The limits of relying on seizure diaries is well documented; however, we chose this outcome measure because it was used in the clinical trials leading to RNS device approval and is the gold standard for judging response for other treatments by the US Food and Drug Administration.^44,45^

There are a number of additional limitations to our study. As a proof-of-principle study, our sample size of 30 patients is small, and gives us limited ability to fully account for center-specific factors related to electrode implantation and non-standardized protocols for RNS programing and outcome reporting across centers. We mitigated variability in data processing by implementing a shared, well-documented pipeline that will easily scale to a larger number of epilepsy centers as we expand patient numbers in future work.^26^ Another limitation is in the retrospective nature of this work. A prospective, randomized clinical trial will ultimately be required to fully assess the benefit of any clinical biomarker or computational model for surgical planning. In this study, we take the first step towards translating our work into clinical practice by preparing a framework to support large-scale validation of our biomarker across centers. Finally, we are limited by the variability in electrode coverage between patients^46^, variability in RNS targets, and heterogeneity in the device settings and medication regimens that may have contributed to responder status.

This study demonstrates a proof-of-principle method for distributed and collaborative RNS biomarker discovery. Future studies will include an expanded number of subjects with additional data containing processed neuroimaging and RNS device recordings. We believe that combining whole-brain structural and functional data along with longitudinal RNS device recordings will be necessary to address major open questions asking who will respond to RNS therapy, where should we implant RNS electrodes, and what are the optimal stimulation parameters for each patient. We hope that our approach and the infrastructure we employ will accelerate the progress of the epilepsy community towards answering these urgent questions.

## Data Availability

We share all functional connectivity networks and synchronizability curves derived from iEEG recordings obtained during a patient's stay in the epilepsy monitoring unit on the free and HIPPA-secure web portal ieeg.org. The code used for generating functional networks and calculating synchronizability is freely available at https://github.com/akhambhati/Echobase.

https://www.ieeg.org

## Abbreviations

AUC: area under the ROC curve
ECoG: electrocorticography
EMU: epilepsy monitoring unit
RNS: responsive neurostimulation
ROC: receiver operating characteristic
SOZ: seizure onset zone

## Acknowledgements

The authors would like to acknowledge Ms. Jacqueline Boccanfuso for her technical assistance with www.ieeg.org.

## References

1. Kwan P, Schachter SC, Brodie MJ. Drug-Resistant Epilepsy.; 2011.

2. Ma BB, Rao VR. Responsive neurostimulation: Candidates and considerations. Epilepsy Behav. 2018;88:388–395. doi:10.1016/j.yebeh.2018.09.032

3. DiLorenzo DJ, Mangubat EZ, Rossi MA, Byrne RW. Chronic unlimited recording electrocorticography–guided resective epilepsy surgery: technology-enabled enhanced fidelity in seizure focus localization with improved surgical efficacy. J Neurosurg. 2014;120(6):1402–1414. doi:10.3171/2014.1.jns131592

4. Nair DR, Laxer KD, Weber PB, et al. Nine-year prospective efficacy and safety of brain-responsive neurostimulation for focal epilepsy. Neurology. 2020;95(9):e1244–e1256. doi:10.1212/WNL.0000000000010154

5. Wei Z, Gordon CR, Bergey GK, Sacks JM, Anderson WS. Implant site infection and bone flap osteomyelitis associated with the NeuroPace responsive neurostimulation system. World Neurosurg. 2016;88:687.e1-687.e6. doi:10.1016/j.wneu.2015.11.106

6. Morrell MJ. Responsive cortical stimulation for the treatment of medically intractable partial epilepsy. Neurology. 2011;77(13):1295–1304. doi:10.1212/WNL.0b013e3182302056

7. Weber PB, Kapur R, Gwinn RP, Zimmerman RS, Courtney TA, Morrell MJ. Infection and erosion rates in trials of a cranially implanted neurostimulator do not increase with subsequent neurostimulator placements. Stereotact Funct Neurosurg. 2017;95(5):325–329. doi:10.1159/000479288

8. Heck CN, King-Stephens D, Massey AD, et al. Two-year seizure reduction in adults with medically intractable partial onset epilepsy treated with responsive neurostimulation: Final results of the RNS System Pivotal trial. Epilepsia. 2014;55(3):432–441. doi:10.1111/epi.12534

9. Nogueira MH, Yasuda CL, Coan AC, Kanner AM, Cendes F. Concurrent mood and anxiety disorders are associated with pharmacoresistant seizures in patients with MTLE. Epilepsia. 2017;58(7):1268–1276. doi:10.1111/epi.13781

10. Jones JE, Hermann BP, Barry JJ, Gilliam F, Kanner AM, Meador KJ. Clinical assessment of axis I psychiatric morbidity in chronic epilepsy: A multicenter investigation. J Neuropsychiatry Clin Neurosci. 2005;17(2):172–179. doi:10.1176/jnp.17.2.172

11. Cendejas Zaragoza L, Byrne RW, Rossi MA. Pre-implant modeling of depth lead placement in white matter for maximizing the extent of cortical activation during direct neurostimulation therapy. Neurol Res. 2017;39(3):198–211. doi:10.1080/01616412.2016.1266429

12. Kokkinos V, Sisterson ND, Wozny TA, Richardson RM. Association of closed-loop brain stimulation neurophysiological features with seizure control among patients with focal epilepsy. JAMA Neurol. 2019;76(7):800–808. doi:10.1001/jamaneurol.2019.0658

13. Chiang S, Khambhati AN, Wang ET, Vannucci M, Chang EF, Rao VR. Evidence of state-dependence in the effectiveness of responsive neurostimulation for seizure modulation. September 2020.

14. Sohal VS, Sun FT. Responsive neurostimulation suppresses synchronized cortical rhythms in patients with epilepsy. Neurosurg Clin N Am. 2011;22(4):481–488. doi:10.1016/j.nec.2011.07.007

15. Liu C, Wen XW, Ge Y, et al. Responsive neurostimulation for the treatment of medically intractable epilepsy. Brain Res Bull. 2013;97(6):39–47. doi:10.1016/j.brainresbull.2013.05.010

16. Razavi B, Rao VR, Lin C, et al. Real-world experience with direct brain-responsive neurostimulation for focal onset seizures. Epilepsia. 2020;61(8):1749–1757. doi:10.1111/epi.16593

17. Arcot Desai S, Tcheng TK, Morrell MJ. Quantitative electrocorticographic biomarkers of clinical outcomes in mesial temporal lobe epileptic patients treated with the RNS® system. Clin Neurophysiol. 2019;130(8):1364–1374. doi:10.1016/j.clinph.2019.05.017

18. Kini LG, Bernabei JM, Mikhail F, et al. Virtual resection predicts surgical outcome for drug-resistant epilepsy. Brain. 2019;142(12):3892–3905. doi:10.1093/brain/awz303

19. Andrews JP, Gummadavelli A, Farooque P, et al. Association of seizure spread with surgical failure in epilepsy. JAMA Neurol. 2019;76(4):462–469. doi:10.1001/jamaneurol.2018.4316

20. Scheid BH, Ashourvan A, Stiso J, et al. Time-evolving controllability of effective connectivity networks during seizure progression. 2020.

21. Proix T, Bartolomei F, Guye M, Jirsa VK. Individual brain structure and modelling predict seizure propagation. Brain. 2017;140(3):641–654. doi:10.1093/brain/awx004

22. Keller CJ, Honey CJ, Mégevand P, Entz L, Ulbert I, Mehta AD. Mapping human brain networks withcortico-ortical evoked potentials. Philos Trans R Soc B Biol Sci. 2014;369(1653). doi:10.1098/rstb.2013.0528

23. Enatsu R, Piao Z, O’Connor T, et al. Cortical excitability varies upon ictal onset patterns in neocortical epilepsy: A cortico-cortical evoked potential study. Clin Neurophysiol. 2012;123(2):252–260. doi:10.1016/j.clinph.2011.06.030

24. Stam CJ, Reijneveld JC. Graph theoretical analysis of complex networks in the brain. Nonlinear Biomed Phys. 2007;1. doi:10.1186/1753-4631-1-3

25. Pecora LM, Carroll TL. Master stability functions for synchronized coupled systems. Phys Rev Lett. 1998;80(10):2109–2112. doi:10.1103/PhysRevLett.80.2109

26. Khambhati AN, Davis KA, Lucas TH, Litt B, Bassett DS. Virtual cortical resection reveals push-pull network control preceding seizure evolution. Neuron. 2016;91(5):1170–1182. doi:10.1016/j.neuron.2016.07.039

27. Litt B, Esteller R, Echauz J, et al. Epileptic seizures may begin hours in advance of clinical onset: A report of five patients. Neuron. 2001;30(1):51–64. doi:10.1016/S0896-6273(01)00262-8

28. Kramer MA, Eden UT, Kolaczyk ED, Zepeda R, Eskandar EN, Cash SS. Coalescence and fragmentation of cortical networks during focal seizures. J Neurosci. 2010;30(30):10076–10085. doi:10.1523/JNEUROSCI.6309-09.2010

29. Fries P. Rhythms for cognition: communication through coherence. Neuron. 2015;88(1):220–235. doi:10.1016/j.neuron.2015.09.034

30. Gómez S, Díaz-Guilera A, Gómez-Gardeñes J, Pérez-Vicente CJ, Moreno Y, Arenas A. Diffusion dynamics on multiplex networks. Phys Rev Lett. 2013;110(2):1–6. doi:10.1103/PhysRevLett.110.028701

31. Wagenaar JB, Brinkmann BH, Ives Z, Worrell GA, Litt B. A multimodal platform for cloud-based collaborative research. In: 2013 6th International IEEE/EMBS Conference on Neural Engineering (NER). IEEE; 2013:1386–1389. doi:10.1109/NER.2013.6696201

32. Ramsay J, Silverman B. Functional Data Analysis. 1st ed. Springer; 1997.

33. Solé-Ribalta A, De Domenico M, Kouvaris NE, Díaz-Guilera A, Gómez S, Arenas A. Spectral properties of the Laplacian of multiplex networks. Phys Rev E. 2013;88(3):032807. doi:10.1103/PhysRevE.88.032807

34. Spencer SS. Neural networks in human epilepsy: evidence of and implications for treatment. Epilepsia. 2002;43(3):219–227. doi:10.1046/j.1528-1157.2002.26901.x

35. Jirsa VK, Proix T, Perdikis D, et al. The Virtual Epileptic Patient: Individualized whole-brain models of epilepsy spread. Neuroimage. 2017;145:377–388. doi:10.1016/j.neuroimage.2016.04.049

36. Goodfellow M, Rummel C, Abela E, Richardson MP, Schindler K, Terry JR. Estimation of brain network ictogenicity predicts outcome from epilepsy surgery. Sci Rep. 2016;6(April):1–13. doi:10.1038/srep29215

37. Kundu B, Davis TS, Philip B, et al. A systematic exploration of parameters affecting evoked intracranial potentials in patients with epilepsy. Brain Stimul. 2020;13(5):1232–1244. doi:10.1016/j.brs.2020.06.002

38. Skarpaas TL, Tcheng TK, Morrell MJ. Clinical and electrocorticographic response to antiepileptic drugs in patients treated with responsive stimulation. Epilepsy Behav. 2018;83:192–200. doi:10.1016/j.yebeh.2018.04.003

39. Deuschl G, Schade-Brittinger C, Krack P, et al. A randomized trial of deep-brain stimulation for Parkinson’s disease. N Engl J Med. 2006;355(9):896–908. doi:10.1056/NEJMoa060281

40. Dell KL, Cook MJ, Maturana MI. Deep Brain Stimulation for Epilepsy: Biomarkers for Optimization. Curr Treat Options Neurol. 2019;21(10):47. doi:10.1007/s11940-019-0590-1

41. Sobayo T, Mogul DJ. Should stimulation parameters be individualized to stop seizures: Evidence in support of this approach. Epilepsia. 2016;57(1):131–140. doi:10.1111/epi.13259

42. Quigg M, Skarpaas TL, Spencer DC, Fountain NB, Jarosiewicz B, Morrell MJ. Electrocorticographic events from long-term ambulatory brain recordings can potentially supplement seizure diaries. Epilepsy Res. 2020;161(October 2019):106302. doi:10.1016/j.eplepsyres.2020.106302

43. Baud MO, Kleen JK, Mirro EA, et al. Multi-day rhythms modulate seizure risk in epilepsy. Nat Commun. 2018;9(1):88. doi:10.1038/s41467-017-02577-y

44. Fisher RS, Blum DE, DiVentura B, et al. Seizure diaries for clinical research and practice: limitations and future prospects. Epilepsy Behav. 2012;24(3):304–310. doi:10.1016/j.yebeh.2012.04.128

45. Cook MJ, O’Brien TJ, Berkovic SF, et al. Prediction of seizure likelihood with a long-term, implanted seizure advisory system in patients with drug-resistant epilepsy: A first-in-man study. Lancet Neurol. 2013;12(6):563–571. doi:10.1016/S1474-4422(13)70075-9

46. Bernabei JM, Arnold TC, Shah P, Revell A, Ong IZ. Electrocorticography and Stereo EEG provide distinct measures of brain connectivity : Implications for network models. MedRxiv. 2020. doi:10.1101/2020.12.02.20242669

