## Supplementary Material for "Synchronizability predicts effective responsive neurostimulation for epilepsy prior to treatment"

### S1. Supplemental Figures and Tables

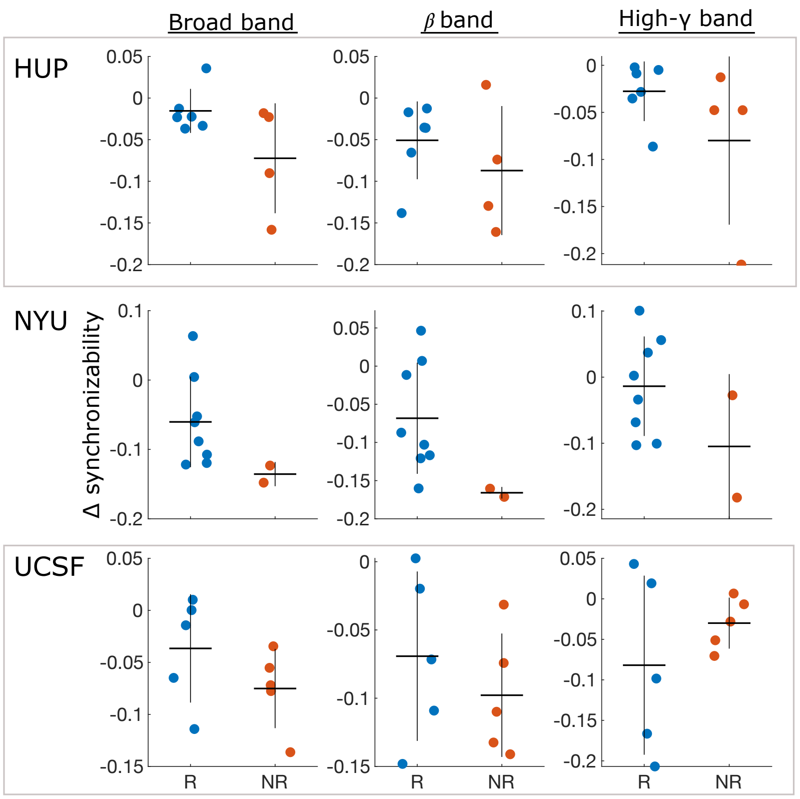


**Figure S1. Center-Specific Synchronizability Analysis.** We calculated the median change in synchronizability after seizure onset for responders (blue points) and non-responders (red points) within each participating epilepsy center. Group level synchronizability trends followed the main results in broad band and $\beta$ band across centers, though no single center reached statistical significance. Heterogeneity in patient trends between centers is evident in the high-γ results. Vertical lines represent the standard deviation within a group and horizontal lines represent the mean. (R- responders, NR- non-responders, HUP- Hospital of the University of Pennsylvania, NYU- New York University Langone, UCSF- University of California, San Francisco Medical Center).

**Table S1. Patient Information.**

| ID | Sex | Age at Onset of Epilepsy | Age at RNS Implant | Baseline Sz/Month | RNS Lead Laterality (R- right, L- left, B- bilateral) | Sz Reduction at Last Visit (%) | Responder Type (R- Responder, NR- Non-Responder) |
|---|---|---|---|---|---|---|---|
| **NP26** | F | 10-19 | 40-49 | 11 | R | 15 | NR |
| **NP28** | M | 30-39 | 30-39 | 225 | L | 17 | NR |
| **NP31** | M | 0-9 | 20-29 | 4 | L | 0 | NR |
| **NP33** | F | 0-9 | 20-29 | 26 | R | 0 | NR |
| **NP35** | F | 20-29 | 30-29 | 53 | L | 71 | R |
| **NP37** | F | 10-19 | 20-29 | 3 | R | 100 | R |
| **NP38** | M | 40-49 | 50-9 | 5 | B | 87 | R |
| **NP40** | M | 20-29 | 40-49 | 53 | L | 29 | NR |
| **NP44** | M | 10-19 | 50-59 | 53 | L | 100 | R |
| **NP47** | M | 0-9 | 20-29 | 30 | B | 100 | R |
| **HUP084** | M | 0-9 | 50-59 | 19 | B | 75-89 | R |
| **HUP108** | M | 10-19 | 30-39 | 13 | B | 1-24 | NR |
| **HUP121** | F | 0-9 | 50-59 | 350 | L | 50-74 | R |
| **HUP127** | F | 10-19 | 30-39 | 19 | B | 0 | NR |
| **HUP128** | F | 10-19 | 60-69 | 64 | L | 100 | R |
| **HUP131** | M | 0-9 | 20-29 | 131 | L | < 0 | NR |
| **HUP137** | M | 30-39 | 50-59 | 24 | B | 75-89 | R |
| **HUP147** | F | 10-19 | 40-49 | 105 | L | 1-24 | NR |
| **HUP153** | F | 40-49 | 50-59 | 4 | B | 50-74 | R |
| **HUP156** | F | 10-19 | 40-49 | 2 | L | 100 | R |
| **NYU01** | F | 0-9 | 20-29 | 5 | L | 88 | R |
| **NYU02** | M | 10-19 | 20-29 | 26 | B | 50 | R |
| **NYU03** | F | 10-19 | 30-39 | 8 | L | 66 | R |
| **NYU04** | M | 20-29 | 40-49 | 5 | B | 100 | R |
| **NYU05** | M | 0-9 | 20-29 | 210 | L | 25 | NR |
| **NYU06** | F | 20-29 | 30-39 | 30 | B | 87 | R |
| **NYU07** | M | 0-9 | 20-29 | 23 | R | 65 | R |
| **NYU08** | M | 10-19 | 20-29 | 6 | B | 48 | NR |
| **NYU09** | F | 0-9 | 40-49 | 53 | L | 95 | R |
| **NYU10** | M | 30-39 | 40-49 | 46 | R | 93 | R |

Sz = seizure.
